# SYSTEMI - Systemic organ communication in STEMI: Design and rationale of a cohort study of patients with ST-segment elevation myocardial infarction

**DOI:** 10.1101/2023.01.13.23284541

**Authors:** F Bönner, C Jung, A Polzin, R Erkens, L Dannenberg, R Ipek, M Kaldirim, M Cramer, P Wischmann, O-P Zaharia, C Meyer, U Flögel, B Levkau, A Gödecke, JW Fischer, N Klöcker, M Krüger, M Roden, M Kelm

**Affiliations:** Department of Cardiology, Pulmonology, and Vascular Medicine, Medical Faculty of Heinrich Heine University, University Hospital, Düsseldorf, Germany; Department of Endocrinology and Diabetology, Medical Faculty of Heinrich Heine University, University Hospital Düsseldorf, Germany; Institute for Clinical Diabetology, German Diabetes Center, Leibniz, Center for Diabetes Research, Düsseldorf, Germany; German Center for Diabetes Research, Partner Düsseldorf, Germany; Departmentn of Cardiology, Evangelisches Krankenhaus, Düsseldorf, Düsseldorf, Germany; Experimental Cardiovascular Imaging, Department of Molecular, Cardiology, Heinrich Heine University, Düsseldorf, Germany; Cardiovascular Research Institute Düsseldorf (CARID), Medical, Faculty, Heinrich Heine University, Düsseldorf, Germany; Institute for Molecular Medicine III, Heinrich Heine University, Düsseldorf, Germany; Institute for Cardiovascular Physiology, Heinrich Heine University, Düsseldorf, Germany; Institute for Pharmacology and Clinical Pharmacology, Heinrich, Heine University, Düsseldorf, Germany; Institute of Neural and Sensory Physiology, Medical Faculty, University of Düsseldorf, Düsseldorf, Germany

**Keywords:** Master Switches, Metabolism, Myocardial ischemia, STEMI

## Abstract

**Background:** (335/350) ST-segment elevation myocardial infarction (STEMI) still causes significant mortality and morbidity despite best-practice revascularization and adjunct medical strategies. Within the STEMI population, there is a spectrum of higher and lower risk patients with respect to major adverse cardiovascular and cerebral events (MACCE) or re-hospitalization due to heart failure. Myocardial and systemic metabolic disorders modulate patient risk in STEMI. Systematic cardiocirculatory and metabolic phenotyping to assess the bidirectional interaction of cardiac and systemic metabolism in myocardial ischemia is lacking.

**Methods:** Systemic organ communication in STEMI (SYSTEMI) is an all-comer open-end prospective study in STEMI patients >18 years of age to assess the interaction of cardiac and systemic metabolism in STEMI by systematically collecting data on a regional and systemic level. Primary endpoint will be myocardial function, left ventricular remodelling, myocardial texture and coronary patency at 6 month after STEMI. Secondary endpoint will be all-cause death, MACCE, and re-hospitalisation due to heart failure or revascularisation assessed 12 month after STEMI. The objective of SYSTEMI is to identify metabolic systemic and myocardial master switches that determine primary and secondary endpoints. In SYSTEMI 150-200 patients are expected to be recruited per year. Patient data will be collected at the index event, within 24 hours, 5 days as well as 6 and 12 months after STEMI. Data acquisition will be performed in multilayer approaches. Myocardial function will be assessed by using serial cardiac imaging with cineventriculography, echocardiography and cardiovascular magnetic resonance. Myocardial metabolism will be analysed by multi-nuclei magnetic resonance spectroscopy. Systemic metabolism will be approached by serial liquid biopsies and analysed with respect to glucose and lipid metabolism as well as oxygen transport. In summary, SYSTEMI enables a comprehensive data analysis on the levels of organ structure and function alongside hemodynamic, genomic and transcriptomic information to assess cardiac and systemic metabolism.

**Discussion:** SYSTEMI aims to identify novel metabolic patterns and master-switches in the interaction of cardiac and systemic metabolism to improve diagnostic and therapeutic algorithms in myocardial ischemia for patient-risk assessment and tailored therapy.

**Trial registration:** **Trial Registration Number:** NCT03539133; **Registration Date** 29.05.2018

**Administrative information:** Note: the numbers in curly brackets in this protocol refer to SPIRIT checklist item numbers. The order of the items has been modified to group similar items (see http://www.equator-network.org/reporting-guidelines/spirit-2013-statement-defining-standard-protocol-items-for-clinical-trials/).

**Funding {4}:** This trial was supported by the German Research Foundation SFB 1116 Grant No. 236177352, as well as project grants BO 4264/1-1 (F.B.); the German Diabetes Center (DDZ), which is funded by the German Federal Ministry of Health and the Ministry of Culture and Science of the state North Rhine-Westphalia and from the German Federal Ministry of Education and Research (BMBF) to the German Center for Diabetes Research (DZD).

***Trials* structured Study Protocol template:** *Trials* guidance: the numbers in curly brackets (e.g. {5a}) are SPIRIT item identifiers. **Please do not remove the numbers in curly brackets, or any heading that contains them.** The item identifiers are slightly out of sequence to make the document flow more easily but it is important that they remain in the document to allow electronic searches by SPIRIT item number. https://trialsjournal.biomedcentral.com/submission-guidelines/preparing-your-manuscript/study-protocoll

## Introduction

### Background and rationale {6a}

ST-segment elevation myocardial infarction (STEMI) continues to confer a substantial burden of morbidity and mortality with plateauing numbers since 2008 [1, 2]. Latecomer STEMI patients, increased patient age and the associated multimorbidity, and STEMI complicated by cardiogenic shock (CS) counterbalance the improvements in prehospital care, ambulance logistics, pharmacotherapy and timely primary percutaneous coronary intervention (pPCI) [2, 3]. Coronary vessel occlusion by plaque erosion and microembolization have become increasingly frequent compared to plaque rupture, which in turn leads to specific types of myocardial ischemia and infarction [4, 5]. At present, almost one-half of patients still demonstrate left ventricular (LV) postinfarct remodeling [6]. Although there is no difference in long-term survival between remodelers and non-remodelers, LV remodelers experience a higher rate of hospitalization for heart failure [6]. This formulates the need to intensify stratification and therapeutic strategies starting in the acute phase of myocardial ischemia [7].

In elderly STEMI patients, metabolic comorbidities such as type 2 diabetes mellitus (T2DM), anemia, and chronic kidney disease (CKD) become increasingly prevalent. Persons with T2DM exhibit a higher incidence of STEMI and an increased mortality risk [8, 9]. When anemia is added to CKD and T2DM, the prognosis further deteriorates [10]. Of note, T2DM is associated with impaired mitochondrial efficacy not only in skeletal muscle [11] but also in cardiomyocytes [12].

Recently, endotypes (subtypes, clusters) of T2DM with distinct metabolic and inflammatory features have been shown to have adverse cardiovascular risk profiles, particularly among patients with a higher degree of insulin resistance [13, 14]. In parallel, the proportion of STEMI patients without standard modifiable cardiovascular risk factors (SMuRFs) has steadily increased from 14 to 27% in the last decade, particularly among younger individuals, with an almost 50% higher 30-day mortality rate than patients with SMuRFs [15]. This highlights the unmet need to identify unknown confounders in the interactions of systemic and cardiac metabolism. In this regard, systematic cardiocirculatory and metabolic phenotyping to assess the bidirectional interaction of cardiac and systemic metabolism in STEMI and during cardiac healing is lacking.

## Objectives {7}

The **objective** of the present cohort study (systemic organ communication in STEMI, SYSTEMI) is to identify metabolic systemic and myocardial master switches that determine outcomes after STEMI.

The aims are as follows: **i)** to characterize myocardial and systemic metabolism in STEMI together with early regional LV function, LV remodeling, myocardial texture and infarct characteristics at index hospitalization **ii)** to characterize the interaction of systemic and myocardial master switches using unsupervised cluster analysis to identify patients with a specific metabolic risk phenotype at index hospitalization **iii)** to identify novel therapeutic master switches and networks prone to interventions and modulations

## Trial design {8}

SYSTEMI is an investigator-initiated, open-end prospective, observational, multicentric, cohort study of all-comer patients with STEMI supported by the German Research Council (CRC 1116, Grant No. 236177352).**The study flow is given in figure 1**.

**Figure 1.**
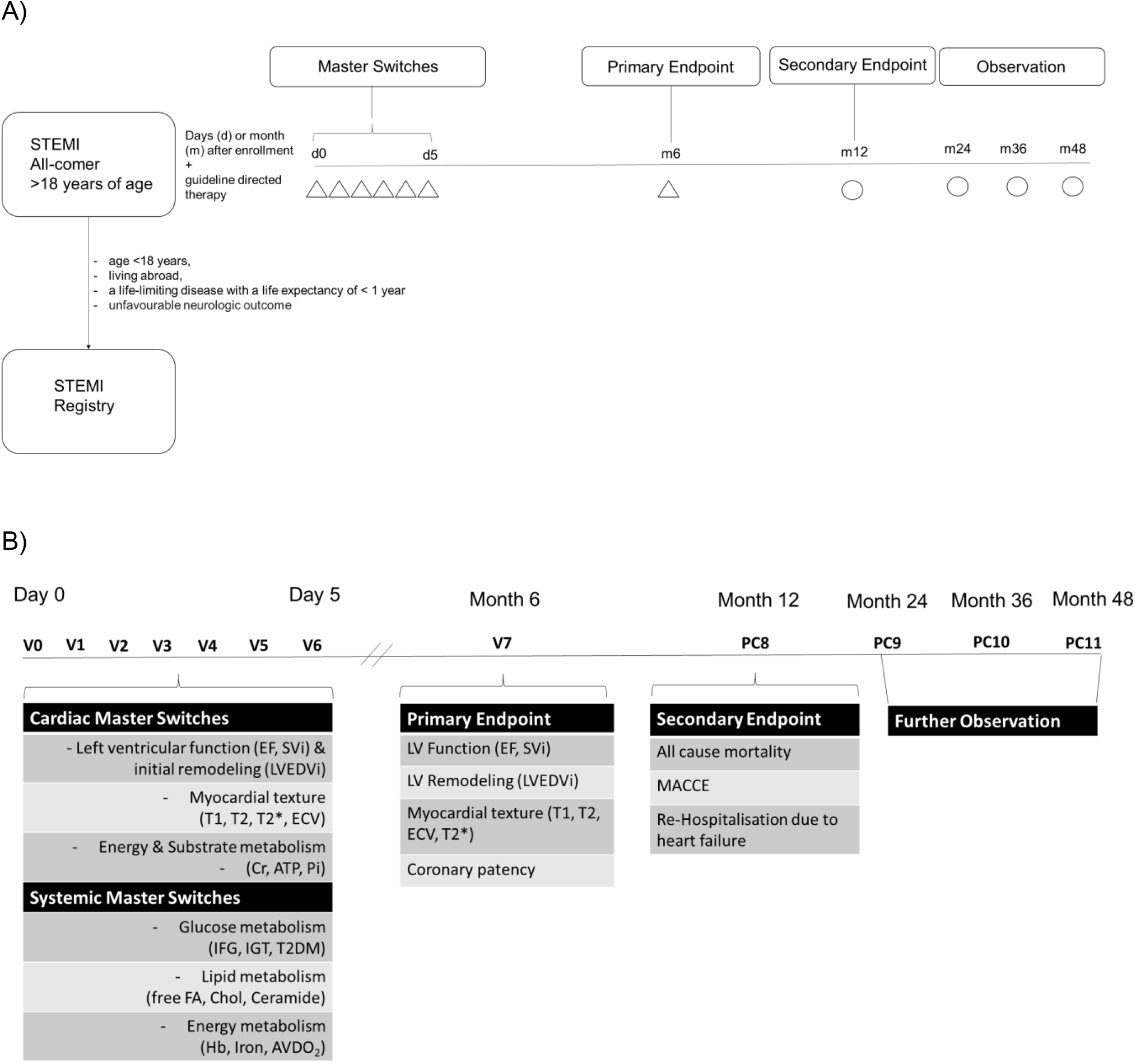
SYSTEMI, systemic organ communication in STEMI, was designed to identify novel metabolic systemic and cardiac masterswitches and networks that determine primary and secondary endpoints, ultimately to improve STEMI care in patient with concomitant metabolic disorders. A) Study Flow Chart of SYSTEMI. Patients with STEMI > 18 years of age will be screened and enrolled for study participation if exclusion criteria are not met. Patients fulfilling exclusion criteria are documented within a separate registry. During index hospitalisation 6 Visits () will be conducted for assessment of myocardial and systemic master switches. At 6 month visit 7 will take place for assessment of the primary endpoints. At 12 month secondary endpoint assessment will be conducted via telephone call while further observation will be conducted yearly. B) Definition of master switches and endpoints. STEMI=ST-segment elevation myocardial infarction, EF=Ejection fraction, SVi=Stroke volume index, WT=Wall thickening, T1=T1-relaxation time, T2=T2-relaxation time, ECV=Extracellular volume fraction, Cr=Creatinine, ATP=Adenosine triphosphate, IFG=Impaired Fasting Glucose, IGT=Impaired Glucose tolerance, T2DM=Type 2 Diabetes Mellitus, FA= Fatty acids, Cho=Cholesterine, Hb=Hemoglobin, AVDO2=arterio-venouse O2 difference, MACCE=Major adverse cardiovascular and cerebral events.

## Methods: Participants, interventions and outcomes

### Study setting {9}

SYSTEMI will recruit patients at the academic main-hospital of Heinrich Heine University Düsseldorf, Germany with >150 STEMI per year, as well as in secondary pPCI -centres within the academic network of Heinrich Heine University Düsseldorf (“Heart-net”), Germany with >80 STEMI per year.

### Eligibility criteria {10}

According to our all-comer design, all patients referred to our hospital with a STEMI diagnosis will be asked for informed consent, if no exclusion criterion is present. The main exclusion criteria are age <18 years, living abroad, and a life-limiting disease with a life expectancy of < 1 year or an unfavourable neurologic outcome. Patients who meet the exclusion criteria will be enrolled in a parallel STEMI register.

### Who will take informed consent? {26a}

Patient selection and short informed consenting will be done by the physician on call at the coronary care unit immediately before the start of pPCI. Before the final consent is given, the investigator or his/her representative will explain verbally the aim, method, source of funding, and the anticipated benefits and potential risks to the participants and answer all questions regarding the study.

### Additional consent provisions for collection and use of participant data and biological specimens {26b}

As part of the general informed consent, all screening subjects are being asked for serial collection of blood samples and its storage in a biobank as well as further genetic analysis. Apart from liquid specimen, electronic patient data are collected systematically and online within the **D**uesseldof **O**utcome, **S**afety and Risk **A**ssessment **R**egistry (**DOSAR**) via health level (HL)-7 interfaces connected to clinical data systems. All requirements of the General Data Protection Regulation are met. A unique participant number will be allocated to each participant (pseudonymization) and assigned chronologically prior to proceeding with study screening. The sequential identification numbers will be used to collect, store, and report participant information.

## Interventions

### Explanation for the choice of comparators {6b}

The treatment of STEMI patient will be conducted according to current guidelines [16]. Details can be seen in SPIRIT Table 1. For assessing initial effects of STEMI on the heart and circulation, we will conduct biplane cineventriculography (CVG) as well as right heart catheterisation during pPCI (V0). This will provide information on LV function and circulatory regulation before reperfusion. During right heart catheterization we will measure cardiac output according to the fick’s law, systemic vascular and pulmonary vascular resistance and AVDO_2_ for oxygen consumption. In parallel, the first liquid biopsies will be generated. The liquid biopsies will serve for assessing basic laboratory parameters including lactate to assess cardiogenic shock and biobanking specimen aiming at subsequent cellular, biochemical, immunological and genetic analyses. Guideline directed repetitive ECG recordings and monitoring will be conducted (V1).

**Table 1.** SPIRIT (Standard Protocol Items: Recommendations for Interventional Trials) figure showing important events and their respective time points during the study period in the cohort study. PE=Patient Empowerment, CA=Coronary Angiography, LHC=Left Heart Catheterisation, RHC=Right Heart atheterisation, CVG=Cineventriculography, TTE=Transthoracic echocardiography, CMR=Cardiovascular Magnetic Resonance, MRS=Magnetic Resonance spectroscopy, FMD=Flow Mediated Dillatation, ECG=Electrocardiogram, OGTT=oral glucose tolerance test, P = Primary Endpoint, S = Secondary Endpoint, O = Observational

| Action | Study Period |  |  |  |  |  |  |  |  |  |  |  |
| --- | --- | --- | --- | --- | --- | --- | --- | --- | --- | --- | --- | --- |
|  | Enrolment | Observation |  |  |  |  |  |  |  |  |  |  |
|  |  | Index Hospitalisation |  |  |  |  |  | FU Hospitalisation | FU Call |  |  |  |
| Visit (V) / Phone Call (PC) | Screening | V1 | V2 | V3 | V4 | V5 | V6 | V7 | PC8 | PC9 | PC10 | P111 |
| Time Point (d) | 0 | 0 | 1 | 2 | 3 | 4 | 5 | 180 | 365 | 730 | 1095 | 1460 |
| *Enrolment* |  |  |  |  |  |  |  |  |  |  |  |  |
| Eligibility Screen | X |  |  |  |  |  |  |  |  |  |  |  |
| Informed Consent | X |  |  |  |  |  |  |  |  |  |  |  |
| Pseudonymisation | X |  |  |  |  |  |  |  |  |  |  |  |
| *Assessments* |  |  |  |  |  |  |  |  |  |  |  |  |
| Mail-Reporting (internal) |  | X | X | X | X | X | X | X |  |  |  |  |
| Questionnaires & PE |  |  | X | X | X | X | X | X |  |  |  |  |
| CA |  | X |  |  |  |  |  | X |  |  |  |  |
| LHC/RHC |  | X |  |  |  |  |  | X |  |  |  |  |
| CVG |  | X |  |  |  |  |  | X |  |  |  |  |
| TTE |  |  | X |  | X |  |  | X |  |  |  |  |
| CMR |  |  | X |  | X |  |  | X |  |  |  |  |
| CMR+MRS |  |  |  |  |  |  | X | X |  |  |  |  |
| Tonometry |  |  |  |  |  |  | X | X |  |  |  |  |
| FMD |  |  |  |  |  |  | X | X |  |  |  |  |
| 4D Flow CMR |  |  |  |  |  |  | X | X |  |  |  |  |
| 12-lead ECG |  | X | X | X |  |  | X | X |  |  |  |  |
| Monitoring |  | X | X | X |  |  |  |  |  |  |  |  |
| Blood Sampling |  | X | X |  |  |  | X | X |  |  |  |  |
| OGTT |  |  |  |  |  |  | X | X |  |  |  |  |
| Blood Sampling |  | X | X |  |  |  | X | X |  |  |  |  |
| *Outcome Assessment* |  |  |  |  |  |  |  |  |  |  |  |  |
| Endpoints |  |  |  |  |  |  |  | P | S | O | O | O |

Within the first and third day after reperfusion we will conduct additional cardiac imaging with transthoracic echocardiography and in a subset of patients cardiovascular magnetic resonance (CMR) (1.5 & 3.0T Achieva, PHILIPS Healthcare, Best, Netherlands) including measurements of ventricular mass, volume, function and myocardial relaxometry (V2+V4). Cine imaging will be used to calculate pressure-volume curves by taking phase-to-phase volumes of short-axis slices, blood pressure and left ventricular end-diastolic pressure measurements from acute catheterization measurements [17]. Additionally, native relaxometric mapping of the myocardium (T1, T2, T2*) [18], gadolinium contrast-enhanced imaging and postcontrast relaxometric mapping (T1, extracellular volume fraction, ECV) will be performed to assess the myocardial inflammation/edema extent, extracellular volume, fibrosis and iron content. For this, validated sequences will be used as outlined in **supplemental table 1**. Gadolinium-enhanced images will be used to assess markers of infarcted tissue, such as edema, infarct size (IS), microvascular obstruction (MVO), and intramyocardial hemorrhage (IMH). Certified CMR evaluation software will be used for all analyses (CVI42, Circle Cardiovascular Imaging Inc., Calgary, Alberta, Canada or Sectra Workstation IDS7, Version 19.3.6.3510). This will be accompanied by an additional liquid biopsy. These early measurements after reperfusion will give insight in very early adaptations in ventricular volume and dimensions, which will serve to associate ventricular volume and dimension with systemic blood derived markers. Blood samples will also be collected to assess arterial or venous lactate, hemoglobin A1c (HbA1c), fasting blood glucose and insulin for calculating the Homeostasis Model Assessment indices (HOMA-IR and HOMA-B). Additionally, a detailed assessment of anemia will be performed, including parameters of RBC turnover, formation, (dys)function, and hemolysis. Specifically, we will address absolute and relative iron deficiency (ID), one of the most common reasons of anemia, especially in elderly patients with comorbidities, by measuring free iron, ferritin, transferrin, the soluble transferrin receptor and transferrin saturation in the collected blood samples. This will be done during V2.

During V2-V5 we will additionally empower the patients to stick to the study by providing detailed information about his health status and study specific examination results. Five days after pPCI (V6), we will again apply echocardiography and CMR with the addition of magnetic resonance spectroscopy (MRS). For regional myocardial metabolic profiling, ^1^H and ^31^P-MRS, which are key techniques for measuring fatty acids (FA) and total creatine as well as ATP, Pi and pH, will be integrated into the clinical CMR protocol 5 days and 6 months after STEMI. For ^1^H-MRS, navigator-free metabolite-cycled spectroscopy using a voxel size of approximately 4-6 ml (10×15×35 mm3) and ≈ 144 averages (7:50 minutes) will be applied [19].

jMRUI/AMARES will also be used to analyze ^31^P spectra. Reconstruction will be performed using a customized reconstruction pipeline in ReconFrame (GyroTools LLC, ZulJrich, Switzerland). jMRUI/AMARES will also be used to analyze MR spectra. Simultaneously, we will have information through liquid biopsies on the diabetes endotype in patients with overt diabetes and on glucose tolerance in patients without diabetes by applying oral glucose tolerance test (OGTT). To asses circulatory adaptations of large conductive arteries, we will apply pulse wave velocity measurements by 4-dimensional velocity-encoding in CMR and by carotid-femoral applanation tonometry using Sphygmocor® technology (AtCor Medical, West Ryde, Australia). The central aortic blood pressure, pulse pressure (PP) and augmentation index (AIX) will be assessed.

#### Examinations at 6, 12 month and on a yearly basis

To assess the primary endpoints, that is LV function, LV remodeling, LV metabolism, LV texture and coronary patency, after complete myocardial healing at 6 months after STEMI, we scheduled further examinations (V7). These examinations comprise of a coronary angiography, echocardiography, CMR, OGTT, applanation tonometry and additional liquid biopsies. The secondary endpoints will be assessed by a telephone call at 12 months (PC8). Further observational phone calls (PC9-11) are planned on a yearly basis.

### Intervention description {11a}

n.a. This cohort study was designed to unravel novel master switches and markers of metabolic networks in STEMI patients to further facilitate risk stratification and identify novel therapeutic targets in STEMI. Currently, no intervention is planed.

### Criteria for discontinuing or modifying allocated interventions {11b}

n.a. This cohort study was designed to unravel novel master switches and markers of metabolic networks in STEMI patients to further facilitate risk stratification and identify novel therapeutic targets in STEMI. We will conduct interim analyses to early identify novel metabolic master switches.

### Strategies to improve adherence to interventions {11c}

n.a. As shown in table 1, each patient will be empowered to adhere to the trial examinations and visits to reach a significant follow up rate.

### Relevant concomitant care permitted or prohibited during the trial {11d}

n.a. This cohort study was designed to unravel novel master switches and markers of metabolic networks in STEMI patients to further facilitate risk stratification and identify novel therapeutic targets in STEMI. STEMI is treated according to current guidelines. There will be no pharmacological trials interfering with the metabolic networks after STEMI.

### Provisions for post-trial care {30}

n.a. Since this trail was designed as cohort study to unravel novel master switches and markers of metabolic networks in STEMI patients along guideline directed therapy, no provisions are being appointed.

### Outcomes {12}

The **primary** endpoints are imaging-derived LV ejection fraction (LVEF), indexed LV stoke volume (SVi), increase in LV indexed end-diastolic volume (LVEDVi) and LV texture (T1, T2, ECV) using either 3-dimensional echocardiography or CMR 6 months after STEMI as well as angiography derived coronary patency rate at 6 month after STEMI. **Secondary** endpoint is a composite of all cause mortality, MACCEs (cardiovascular death, nonfatal stroke or myocardial infarction) and rehospitalization due to heart failure within 12 months after STEMI.

### Participant timeline {13}

#### SPIRIT TABLE 1

##### Sample size {14}

SYSTEMI was designed as open-end prospective cohort study in all comer STEMI patients. No formal sample size calculation has been performed. It was anticipated that more than 160 STEMI patients per year will be a realistic recruitment speed on the basis of expected STEMI patients at host institutions to reach a total sample size of >1000 STEMI patients.

##### Recruitment {15}

SYSTEMI will screen every patient suffering from STEMI >18 years of age. Every patient will be given a sequential identification number. To optimize recruitment efficacy, training sessions have been performed with the interventional staff on call and the study nurses to best inform the patients about the study dependent examinations during their guideline directed diagnostics and therapy at index hospitalisation.

## Assignment of interventions: allocation

### Sequence generation {16a}

n.a. SYSTEMI was designed as observational cohort study. Pseudonymisation numbers are generated consecutively.

### Concealment mechanism {16b}

n.a. This cohort study was designed to unravel novel master switches and markers of metabolic networks in STEMI patients to further facilitate risk stratification and identify novel therapeutic targets in STEMI. Pseudonymisation numbers are generated consecutively.

### Implementation {16c}

n.a. SYSTEMI was designed as observational cohort study. Pseudonymisation numbers are generated consecutively

## Assignment of interventions: Blinding

### Who will be blinded {17a}

n.a. SYSTEMI was designed as observational cohort study and thus interventions with necessary blinding operations are not scheduled. Pseudonymisation numbers are generated consecutively.

### Procedure for unblinding if needed {17b}

n.a. SYSTEMI was designed as observational cohort study and thus interventions with necessary blinding operations are not scheduled. Pseudonymisation numbers are generated consecutively.

## Data collection and management

### Plans for assessment and collection of outcomes {18a}

Patient data are collected systematically and online within the DOSAR Registry via health level (HL)-7 interfaces connected to clinical data systems. All requirements of the General Data Protection Regulation are met. The SYSTEMI Steering Committee is responsible for the scientific content of the protocol, oversees the study steps, and checks adherence to “Good Clinical Practice” and the study protocol.

### Plans to promote participant retention and complete follow-up {18b}

As can be seen in table 1, patient empowerment is done on a regular, daily basis by the study team, which consists of the interventionalist, which was given initial informed consent, the physician on call within the STEMI program and the study nurse.

### Data management {19}

Patient data are collected systematically and online within the DOSAR registry via health level (HL)-7 interfaces connected to clinical data systems. All requirements of the General Data Protection Regulation are met.

### Confidentiality {27}

Patient data are collected in a pseudonymised manner. The clinical work-flow of patients will not be intervened.

### Plans for collection, laboratory evaluation and storage of biological specimens for genetic or molecular analysis in this trial/future use {33}

By informed consent, every patient recruited to SYSTEMI is aware of genetic analysis of blood derived biosamples. Biosamle processing and storage is done according to existing standard operating procedures (SOPs) at -80°C. Biosample access is regulated by the SOPs and granted by standardized evaluation upon formal request.

## Statistical methods

### Statistical methods for primary and secondary outcomes {20a}

This cohort study was designed to unravel novel master switches and markers of metabolic networks in STEMI patients to further facilitate risk stratification and identify novel therapeutic targets in STEMI. For primary endpoints, logistic regression analysis will be used to model the predefined outcome data with impact of covariates. The effects of the independent parameters in these regression models will be estimated (odds ratio, hazard ratio) and presented with 95% confidence intervals. Network analysis using machine learning approaches for unsupervised metabolic cluster identification will be conducted. Secondary endpoint analysis will be performed using Kaplan–Meier plots and the log-rank test. Two-tailed p values below 0.05 will be considered indicative of statistically significant differences.

### Interim analyses {21b}

This cohort study was designed to unravel novel master switches and markers of metabolic networks in STEMI patients to further facilitate risk stratification and identify novel therapeutic targets in STEMI. Since significant interactions of master switches within the metabolic network are anticipated, interim analysis will be done after 500 and 1000 patients for a robust statistical power calculation and re-adjustment of investigations.

### Methods for additional analyses (e.g. subgroup analyses) {20b}

There are several possibilities for subgroup analysis depending on specific patient phenotypes along the glucometabolic spectrum. These analyses will be conducted after the interim analysis.

### Methods in analysis to handle protocol non-adherence and any statistical methods to handle missing data {20c}

Protocol non-adherence is documented within DOSAR.

### Plans to give access to the full protocol, participant level-data and statistical code {31c}

Upon formal request according to our internal SOP, access to primary data can be given. This process is guided by the SYSTEMI Steering Committee, which is responsible for the scientific content of the study data.

## Oversight and monitoring

### Composition of the coordinating centre and trial steering committee {5d}

The SYSTEMI Steering Committee is responsible for the scientific content of the protocol, oversees the study steps, and checks adherence to “Good Clinical Practice” and the study protocol as well as performance. Study conduction will follow the SOP of the Clinical Trial Unit (CTU) of the University Hospital of Düsseldorf. The SYSTEMI Endpoint Adjudication Committee (EAC) will adjudicate clinical endpoints based on data provided by the clinical trial sites. This procedure will be supervised by the trial coordination center (TCC, Koordinierungszentrum für Klinische Studien, KKS) at Heinrich Heine University. The trial management committee (TMC) meets once a week and the steering committee twice a year to review trial conduct.

### Composition of the data monitoring committee, its role and reporting structure {21a}

The data monitoring committee (DMC) consist of a data manager and participants of the external advisory board. The DMC will report in a weekly manner organizational aspects of data collection and handling as well as quality and completeness reports.

### Adverse event reporting and harms {22}

n.a. In SYSTEMI there will be no use of medical products. Patient care is guideline directed. Thus, adverse or even serious adverse events are not expected by definition.

### Frequency and plans for auditing trial conduct {23}

Data quality and completeness is of prime importance in SYSTEMI. Regular audits are conducted synchronized to the annual meeting of the CRC 1116 and Cardiovascular Research Institute Düsseldorf (CARID).

### Plans for communicating important protocol amendments to relevant parties (e.g. trial participants, ethical committees) {25}

If applicable, relevant trial modifications will be communicated via trail registration page at ClionicalTrails.gov NCT03539133 and the ethical committee for protocol version update.

## Dissemination plans {31a}

Results of SYSTEMI will be disseminated by publications in peer reviewed journals. Furthermore, SYSTEMI is connected to the annual reporting structure of CARID. Quality reports and interim analysis will be presented on annual CARID conferences.

## Discussion

Mortality and morbidity after STEMI remain high and current risk prediction models do not take metabolic risk constellations into account. Therefore, the overall objective of SYSTEMI is to identify systemic and myocardial metabolic master switches that determine outcomes after STEMI. To reach this objective, we have designed and set-up SYSTEMI to comprehensively investigate systemic and myocardial metabolism at index hospitalization and at 6 months in more than 1000 patients. SYSTEMI will enable in-depth analyses of systemic metabolic effectors on local myocardial function and metabolism, as detected by magnetic resonance imaging and spectroscopy, and clinical outcome.

### SYSTEMI metabolic clusters

To our knowledge, the present cohort study is the first study in patients with STEMI to systematically and simultaneously assess organ-function, inter-organ communication as well as myocardial and systemic metabolism. SYSTEMI focuses on 3 metabolic clusters: i) the glucometabolic spectrum, ii) the lipid metabolism and iii) the O_2_ transport capacity of the blood.

The first metabolic cluster covers the glucometabolic spectrum of hyperglycemia, hyperinsulinemia, and insulin resistance to T2DM and the role of specific endotypes of T2DM in patients with STEMI. Altered glucose metabolism can be seen in up to 50% of all STEMI patients, but only 25% remain in this state after 3 months [20]. Moreover, the hyperglycemic status of diabetic and nondiabetic patients has been shown to influence IS, MVO and LVEF post-STEMI [20]. Therefore, SYSTEMI will focus on glucometabolic phenotyping using various methods, including OGTT, homeostatic model assessment, or endotyping of T2DM. Endotyping of T2DM has shown different prevalence rates of diabetes-related complications, although their impact on STEMI patients remains unknown [13, 14]. In this context, the SIRD endotype may specifically predispose STEMI patients to metabolic and myocardial alterations. SIRD has been associated with elevated levels of biomarkers of inflammation and a higher risk of diabetic kidney disease [13, 14]. This characterization may help to guide targeted therapeutic regimens for patients with STEMI and T2DM. The recent introduction of sodium-glucose cotransporter-2 inhibitors into guidelines for heart failure in patients with and without T2DM has highlighted the importance of glucometabolic modulation in cardiovascular disease to improve patient outcomes [21].

The second metabolic cluster covers the crosstalk of cardiac ischemia with changes in lipid metabolism in the presence and absence of alterations of the glucometabolic spectrum. The impacts of browning adipose tissue [22], the lipolytic response in skeletal muscle, and circulating lipoproteins, ceramides, and the HDL sphingolipidome [23] on infarct will be investigated.

The third metabolic cluster covers function and dysfunction of red blood cells in anemia [24]. Anemia affects systemic energy metabolism through multiple mechanisms, including limited oxygen transport capacity, disturbed iron metabolism, and altered hepatic metabolism, and is associated with profound red blood cell (RBC) dysfunction [24]. This appears to be an important modulator of interorgan crosstalk after STEMI. The prognosis of STEMI is limited by anemia and secondary complications such as bleeding, thrombosis, inflammation, and arrhythmia [25].

### SYSTEMI approaches to characterize myocardial and systemic metabolism

To enable comprehensive investigation of the bidirectional interaction of systemic and cardiac metabolism through the aforementioned pathways in the acute phase of STEMI and its long-term outcome in patients, several layers of analysis have to be accomplished within SYSTEMI: (i) analysis of cardiac and circulatory function prior to and immediately after pPCI, on the 1^st^, 3^rd^, and 5^th^ day of the index hospitalization using novel CMR techniques, (ii) assessment of glucose and lipid metabolism, and (iii) readouts of various organ and tissue functions during and after STEMI, including the liver, the pancreas, the spleen, and circulating blood and immune cells arising from bone marrow, the kidney, skeletal muscle, and adipose tissue. This is complemented by targeted multiomics analysis, including proteomics, lipidomics, and long-read sequencing, and novel CMR-based techniques to study myocardial flux rates and energy metabolism in mice and patients.

In perspective, SYSTEMI seeks to link regional functional and structural alterations with local myocardial metabolism. Several technical improvements in recent years have facilitated the application of MRS, ^1^H- and ^31^P-MRS measurements by reducing the scan time or voxel size for examining metabolism in infarcted and remote zones [19]. In experimental models of myocardial ischemia, substrate selection is characterized by a “metabolic switch” in myocardial energy supply from predominantly aerobic FA oxidation to anaerobic glycolysis. ^1^H-MRS may estimate myocardial triglyceride and total creatine levels, whereas ^31^P-MRS is able to quantify metabolites such as phosphocreatine, ATP, their ratio or creatine kinase fluxes [26]. Currently, data are lacking for CMR/MRS measurements that comprehensively characterize metabolic alterations in ischemic and remote myocardium with respect to coronary artery disease severity using ^1^H- and ^31^P-MRS. In perspective, myocardial metabolism will be further assessed by use of hyperpolarized ^13^C-substrates, such as pyruvate, bicarbonate or lactate, combined with advanced deuterium metabolic imaging [27].

### Conclusion

SYSTEMI aims to identify individuals with distinct susceptibility to myocardial ischemia among the different endotypes of T2DM across the glucometabolic spectrum, endotypes of anemia and lipid disorders, paving the way for precision medicine in STEMI and cardiometabolic disorders.

## Trial status

After study registration, a run in phase was initiated in 2017 to implement clinical and analytical interfaces and to acquire datasets from all clusters with optimal completeness. In January 2018 the first patient was recruited to SYSTEMI according to the protocol version 2.0. The trial is currently running and will be conducted open-end. Interim analysis will be conducted every 500 enrolled patient.

## Data Availability

All data produced in the present work are contained in the manuscript

## Abbreviations

AI: = Artificial intelligence
CEST: = Chemical exchange saturation transfer
CS: = Cardiogenic shock
CKD: = Chronic kidney disease
CMR: = Cardiac magnetic resonance
CRC: = Collaborative Research Consortium
DMC: = Data Monitoring Committee
DOSAR: = Düsseldorf outcome and safety registry
EAC: = Endpoint Adjunction Committee
EF: = Ejection fraction
ECV: = Extracellular volume fraction
EDV: = End-diastolic volume
ESV: = End-systolic volume
FMD: = Flow-mediated dilatation
FU: = Follow-up
HbA1c: = Hemoglobin A1c
HOMA: = Homeostasis model assessment I
MH: = Intramyocardial hemorrhage
ID: = Iron deficiency
IS: = Infarct size
LAS: = Large artery stiffness
LDL-C: = Low-density lipoprotein cholesterol
LV: = Left ventricle
MACEs: = Major adverse cardiovascular events
MRS: = Magnetic resonance spectroscopy
MVO: = Microvascular obstruction
6MWT: = 6 minutes walk test
NAFLD: = Non-alcoholic fatty liver disease
OGTT: = Oral glucose tolerance test
pPCI: = Primary percutaneous coronary intervention
PWV: = Pulse wave velocity
SMuRF: = Standard modifiable cardiovascular risk factor
SOP: = Standard operating procedure
STEMI: = ST-segment elevation myocardial infarction
SV: = Stroke volume
SYSTEMI: = Systemic organ communication in STEMI
TMC: = Trial Management Committee
T2DM: = Type 2 diabetes mellitus

## Declarations

The authors have nothing to declare

## Acknowledgements

Joy Dillenburg, Stefanie Bensmann, Marina Masyuk, Sebastian Haberkorn, Fabian Nienhaus, Juliane Geisler, Elisabetha Gharib, Alicja Timm, Sheila Kendic. Figure 3 was created with BioRender.com.

## Authors’ contributions {31b}

Authors' contributions {31b}
| Author | Study Idea | Study Design | Study Conduction | Interpretation of study data | Drafting the manuscript |
| --- | --- | --- | --- | --- | --- |
| Florian Bönner |  | X | X | X | X |
| Christian Jung |  | X | X | X | X |
| Amin Polzin |  |  | X |  | X |
| Ralf Erkens |  |  | X |  | X |
| Mareike Cramer |  |  | X | X | X |
| Lisa Dannenberg |  |  | X |  | X |
| Rojda Ipek |  |  | X |  | X |
| Madlen Kaldirim |  |  | X |  | X |
| Patricia Wischmann |  |  | X |  | X |
| Patricia Zaharia |  |  | X |  | X |
| Christian Meyer |  |  | X |  | X |
| Ulrich Flögel |  |  |  |  | X |
| Bodo Levkau |  |  |  |  | X |
| Axel Gödecke |  |  |  |  | X |
| Jens Fischer |  |  |  |  | X |
| Martina Krüger |  |  |  |  | X |
| Nico Klöcker |  |  |  |  | X |
| Michael Roden | X | X | X | X | X |
| Malte Kelm | X | X | X | X | X |

## Funding {4}

This trial was supported by the German Research Foundation SFB 1116 Grant No. 236177352, as well as project grants BO 4264/1-1 (F.B.); the German Diabetes Center (DDZ), which is funded by the German Federal Ministry of Health and the Ministry of Culture and Science of the state North Rhine-Westphalia and from the German Federal Ministry of Education and Research (BMBF) to the German Center for Diabetes Research (DZD).

### Availability of data and materials {29}

Data of SYSTEMI will be available after a reasonable request from the SYSTEMI Steering Committee. All data will be curated and organized into a set of files, shared on an online, publicly available data repository after peer-reviewed publication (preservation and accessibility of the data) to ensure potential secondary analyses, long-term archiving, and reuse by other researchers (scientific recognition). Besides the study protocol, publications are planned for the results in peer-reviewed journals. In addition, results will be communicated in lay language to participants and health care providers as they may affect current dietary recommendations for patient with peanut and or tree nut allergy.

### Ethics approval and consent to participate {24}

Ethics approval was given by the local ethics committees (Heinrich-Heine-University, Düsseldorf, Germany: 5961R and 2021055 as well as Landesärztekammer Nordrhein: 2021055) complying with the Declaration of Helsinki. SYSTEMI is registered at www.clinicaltrials.gov: NCT03539133.

## Consent for publication {32}

As this is an investigator-initiated trial, only the principal investigators, sub-investigators, and collaborators are responsible and involved in the publications Authors using data from SYSTEMI are directed to announce the CRC 1116 Grant No. 236177352

## Authors’ information (optional)

*n*.*a*. additional information upon reasonable request

---

**Supplemental Table 1.** CMR sequences used and respective references

| CMR Sequences |  |  |
| --- | --- | --- |
| Item | Technique | Sequence + DOI |
| Volumes, global and local function, myocardial mass | Cine in 4-,3-2ChV, and SA | SSFP<br>repetition time (TR)/echo time (TE) = shortest, flip angle (FA) = 60°, reconstructed voxel size = 1.46 × 1.45 × 8 mm<sup>3</sup>, end-expiratory breath-hold<br>https://doi.org/10.1186/s12968-015-0118-0 |
| Oedema | T2w-imaging<br>T2-mapping | GRaSE<br>TR = 1 RR interval, number of echo images = 15, echo spacing 10 ms, leading to an echo train of 150 ms, number of gradient echoes for segmented acquisition = 3 (EPI factor), FA = 90°, spatial resolution: 2 × 2 × 10 mm<sup>3</sup>, parallel imaging (SENSE) acceleration factor of 2, k-space data acquired with cartesian encoding scheme. Double inversion black-blood pulse<br>https://doi.org/10.1186/s12968-015-0118-0 |
| Haemorrhage | T2*-mapping | FFE<br>TR shortest, TE first: short, number of echo images = 6, echo spacing shortest, FA = 20°, spatial resolution 2 × 2 × 8 mm<sup>3</sup>, parallel imaging (SENSE), fast imaging: TFE, TFE pre-pulse black blood<br>https://doi.org/11.2234/s32345-232-34-432 |
| Fibrosis | T1-mapping<br>ECV-mapping | MOLLI<br>Balanced steady state free precession single breath-hold modified Look-Locker Imaging (MOLLI, (3(3)3(3)5)) (TE/TR/flip-angle: 1.64msec/3.3msec/50°, acquired voxel size 1.8x1.8x8 mm, phase encoding steps n=166, 11 images corresponding to different inversion times (3+3+5 MOLLI scheme), adiabatic prepulse to achieve complete inversion)<br>https://doi.org/10.1016/j.ejrad.2016.10.031. |
| Cardiac and thoracic fat volume | mDIXON | FFE<br>TR shortest, TE first: short, number of echo images = 2, echo spacing shortest, FA = 15°, spatial resolution 1.25 × 1.5 × 1.5 mm<sup>3</sup>, 100 slices, parallel imaging (SENSE), fast imaging: TFE, TFE pre-pulse no<br>https://doi.org/10.1007/s00330-020-07517-x |
| Myocardial triglycerides | <sup>1</sup>H Spectroscopy | PRESS<br>minimum TR 1 beat, TE shortest, FA = 90°, metabolite cycling by adding an optimized Hwang pulse in front of the sequence, bandwidth 2000 Hz (1024 samples), voxel size from 5 x 10 x 25 mm, pencil beam volume shimming during a single breathold, triggering to maximum systole, 144 averages, free breathing acquisition |
| Vascular Function | 4D Flow | FFE<br>TR shortest, TE first: short, number of echo images = 1, FA = 5.2°, spatial resolution 2.8 × 2.8 × 2.8 mm<sup>3</sup>, 40 slices, parallel imaging (SENSE), fast imaging: TFE, TFE pre-pulse no |
| Microvascular patency | Fist Pass Perfusion<br>(gadoteridol, ProHance®,<br>Bracco Imaging, total dose =<br>0.2 mmol/kgKG) | FFE<br>TR shortest, TE shortest, number of echo images =<br>1, FA = 50°, spatial resolution $3 \times 3 \times 10$ mm<sup>3</sup>, 3<br>slices, parallel imaging (SENSE), fast imaging:<br>TFE, TFE pre-pulse saturate<br>https://doi.org/10.1186/s12968-015-0118-0 |
| Infarct Size | Late Gadolinium<br>enhancement (10 Minutes)<br>(gadoteridol, ProHance®,<br>Bracco Imaging, total dose =<br>0.2 mmol/kgKG) | Turbo FFE<br>TR shortest, TE shortest, 3D gradient spoiled, 180°<br>inversion, FA = 15°, °, spatial resolution $1.52 \times$<br>$1.71 \times 10$ mm<sup>3</sup>, reconstructed voxel size = $1.52 \times$<br>$1.52 \times 5$ mm<sup>3</sup>, end-diastolic, end-expiratory breath-hold<br>https://doi.org/10.1186/s12968-015-0118-0 |
| Microvascular Obstruction | Late Gadolinium<br>enhancement (5 & 15<br>Minutes)<br>(gadoteridol, ProHance®,<br>Bracco Imaging, total dose =<br>0.2 mmol/kgKG) | Turbo FFE<br>TR shortest, TE shortest, 3D gradient spoiled, 180°<br>inversion, FA = 15°, °, spatial resolution $1.52 \times$<br>$1.71 \times 10$ mm<sup>3</sup>, reconstructed voxel size = $1.52 \times$<br>$1.52 \times 5$ mm<sup>3</sup>, end-diastolic, end-expiratory breath-hold<br>https://doi.org/10.1186/s12968-015-0118-0 |

